# *DLST*—a Cuproptosis-related Gene—is a Potential Diagnostic and Prognostic Factor for Clear Cell Renal Cell Carcinoma

**DOI:** 10.1101/2023.04.27.23289219

**Authors:** Haoyuan Wang, Xiaopeng Ma, Sijie Li, Xiaochen Ni

## Abstract

Clear cell renal cell carcinoma (ccRCC) accounts for the highest number of renal malignancies and 3% of all adult cancers. The incidence of ccRCC is increasing worldwide, and its prognosis is poor. Approximately 30% of the patients are diagnosed at a late stage and are frequently asymptomatic. Cuproptosis is a new type of cell death that is regulated by Cu ions. As cuproptosis is associated with cancer development, we hypothesized that changes in the expression of cuproptosis-related genes (CRGs) are associated with the prognosis of ccRCC, and that CRGs can serve as biomarkers for the diagnosis and prognosis of ccRCC. In the present study, we explored the correlation between CRGs and ccRCC prognosis by analyzing publicly available data. We analyzed the clinical information and RNA-sequencing data in The Cancer Genome Atlas using bioinformatics tools. Dihydrolipoamide S-succinyltransferase (*DLST*) was identified as a novel gene with predictive and diagnostic potential. CRGs were under-expressed in ccRCC samples, and downregulation of *DLST* was highly associated with poor prognosis. Cox univariate and multivariate regression analyses revealed that *DLST* could serve as an independent prognostic factor for ccRCC. Further, functional enrichment analysis indicated that low expression of *DLST* may affect immune function. Our results strongly indicate that *DLST* plays an important role in ccRCC progression and may serve as an independent diagnostic and prognostic biomarker for ccRCC. Therefore, *DLST* is a potential therapeutic target for patients with ccRCC.

## 1 Introduction

Renal cell carcinoma (RCC) is the most common form of kidney cancer, and clear cell RCC (ccRCC) is one of the most common types of RCC, accounting for 70–90% of the histologic subtypes of RCC (Chen et al. 2019, 2020). ccRCC is the most common malignancy of the urinary system, with more than 400,000 cases and approximately 175,000 deaths reported worldwide every year (Carril-Ajuria et al. 2019). Surgical resection is considered the most effective treatment for RCC because it is resistant to radiotherapy and chemotherapy; however, the outcome remains unsatisfactory in advanced or metastatic cases. Most patients die within 2–3 years of diagnosis, 30% exhibit metastases after initial diagnosis, and approximately 40% patients develop distal metastases after surgery (Xu et al. 2020). Therefore, a better understanding of the pathogenesis of ccRCC and the identification of biomarkers for diagnosis, prognosis, and treatment are imperative.

Cu, an essential trace metal, is required for maintaining body homeostasis and for the metabolic regulation of many cell types (Ruiz et al. 2021). Cu deficiency causes malfunctioning of Cu-binding enzymes, whereas its excess causes cell death (J. Chen et al. 2020). However, Cu is also found in the serum and tumor tissues of patients with breast, lung, colorectal, cervical, and ovarian cancers (Z. Li et al. 2022; Wang et al. 2022; Y. Chen 2022; Xu et al. 2022). Moreover, Cu affects tumor occurrence, invasion, and metastasis. Previous studies have reported that Cu levels are higher in cancer patients than in healthy controls. When Cu accumulates within cells, it binds to the lipid-acylated proteins of the tricarboxylic acid (TCA) cycle in the mitochondria, and Fe–S cluster proteins are lost, causing proteotoxic stress-induced death called “cuproptosis” (Tang et al. 2022; Cobine and Brady 2022). Cuproptosis is a newly discovered form of cell death that differs from apoptosis, ferroptosis, and necroptosis. Cuproptosis involves neither cleavage of caspase-3 activity, a hallmark of apoptosis, nor activation of apoptosis. Treatment with other inhibitors of known cell death mechanisms, including ferrostatin-1, necrostatin-1, and N-acetylcysteine (oxidative stress), failed to abrogate copper ionophore-induced cell death. Several studies have shown that mitochondrial dysfunction contributes to kidney disease, and the inhibition of mitochondrial respiration enhances the efficacy of chemotherapy for RCC. We searched the literature on cuproptosis and selected eight genes that had not yet been identified. Thus, based on previous studies, we verified cuproptosis-related genes (CRGs), namely, dihydrolipoamide S-succinyltransferase (*DLST*), ferredoxin 1 (*FDX1*), pyruvate dehydrogenase complex component X (*PDHX*), cytochrome C oxidase subunit 7B (*COX7B*), solute carrier family 6 member 3 (*SLC6A3*), NADH: ubiquinone oxidoreductase subunit B1 (*NDUFB1*), copper chaperone for superoxide dismutase (*CCS*), and midline 1 (*MID1*), which provide a preliminary basis for this study.

*DLST* is a protein-coding gene and is associated with transferase activity and dihydrolipoyllysine-residue succinyltransferase activity (Anderson et al. 2016; Yang et al. 2009). Lipoylation of these proteins is essential for enzymatic activity, and Cu induces cell death through protein lipoylation. Thus, cells with high levels of lipoylated proteins are susceptible to Cu-induced cell death. Therefore, we hypothesized that the downregulation of *DLST* expression promotes tumor progression by inhibiting cuproptosis in ccRCC cells.

In this study, comprehensive bioinformatic analyses were performed to verify the eight CRGs with diagnostic and prognostic potential in ccRCC. We analyzed the RNA-sequencing (RNA-seq) data downloaded from The Cancer Genome Atlas (TCGA), the Genotype-Tissue Expression (GTEx), and Gene Expression Omnibus (GEO). *DLST* was identified as a novel diagnostic and prognostic biomarker for ccRCC.

## 2 Materials and Methods

### 2.1 Data acquisition

TCGA Kidney Renal Clear Cell Carcinoma (KIRC) RNA-seq data and clinical information, as well as GTEx normal kidney RNA-seq data, were downloaded from UCSC Xena (https://xenabrowser.net/datapages/) (Goldman et al. 2020). We obtained data of 531 ccRCC tissue samples, 100 normal kidney tissue samples, including 28 normal samples, and 72 paracancerous normal tissue samples from TCGA-KIRC, GTEx, TCGA, respectively. All transcript per million (TPM)-formatted RNA-seq data from UCSC Xena were processed by Toil in UCSC Xena. The GSE78179 and GSE53757 datasets were downloaded from the GEO database (https://www.ncbi.nlm.nih.gov/geo/) (Li et al. 2022). GSE78179 contains six samples: three metastatic ccRCC samples and three healthy kidney epithelial tissues (Khan et al. 2016). GSE53757 contains 72 advanced and metastatic ccRCC samples and 72 normal kidney samples (von Roemeling et al. 2014). Eight CRGs (*DLST, FDX1, PDHX, COX7B, SLC6A3, NDUFB1, CCS*, and *MID1*) were identified from the literature. DLST immunohistochemical profiles of normal kidney and ccRCC tissue samples were obtained from the Human Protein Atlas (www.proteinatlas.org) (Han et al. 2020).

### 2.2 Differential expression analysis of CRGs

For differential expression analysis, we identified eight differentially expressed CRGs between ccRCC and normal samples from TCGA-KIRC and GTEx data. The changes in expression between the ccRCC samples and adjacent normal tissues were calculated using the “DESeq2” R package; we also calculated the log_2_ fold change (FC) and p-value adjusted by the FDR. The difference ordering diagram was constructed based on the size of the final log_2_ FC to exhibit the differential expression, and the CRGs were marked in the diagram. Boxplot graphs were created using the “ggplot2” R package. The GSE78179 and GSE53757 datasets were used to validate the differential expression of CRGs. Heat maps were constructed using the “ComplexHeatmap” R package (https://github.com/jokergoo/ComplexHeatmap/). To avoid the confounding effects of different histological origins, we selected 72 paired samples from TCGA. Differences were considered statistically significant when |log_2_ FC| > 1 and adjusted p < 0.05.

### 2.3 Correlation analysis and construction of gene–gene and protein–protein interaction networks

The correlations among the eight CRGs in TCGA-KIRC dataset were analyzed using Spearman correlation analysis. A correlation heat map was generated using the “ggplot2” R package. Gene networks were visualized, and gene functions were predicted using GeneMANIA (https://genemania.org/) (Warde-Farley et al. 2010). We constructed a network based on co-expression, and predicted colocalization, genetic interactions, and shared protein domains in the GeneMANIA database (https://genemania.org/). STRING (https://cn.string-db.org/) was used to predict the potential interactions between the proteins encoded by the eight CRGs (He et al. 2020).

### 2.4 Diagnostic and prognostic value of eight CRGs in ccRCC

The clinical information in TCGA dataset comprises age, sex, race, TNM stage, pathologic stage, primary therapy outcome, histologic grade, serum calcium level, hemoglobin level, laterality, overall survival (OS) event, disease-specific survival (DSS) event, and progression-free interval (PFI) event (Table 1). A receiver operating characteristic (ROC) curve was developed to analyze the predictive efficacy of the expression of the eight CRGs in tumor appearance, using the “pROC” package in R. The Kaplan–Meier survival curves of TCGA data were used for survival analysis between the samples with low and high expression levels of the eight CRGs, using the “survival” package in R. We then selected the prognostic genes using univariate Cox regression analyses. Based on a forest plot made using “ggplot2,” the hazard ratio (HR) and p-value for all eight CRGs were calculated. An independent prognostic factor was identified using the Cox proportional hazard regression analysis. Next, we assessed the prognostic relevance of *DLST* expression in predicting OS and disease-free survival (DFS) of ccRCC patients, using the GEPIA2 data portal (http://gepia2.cancer-pku.cn) (Tang et al. 2019). Using boxplots, we also compared the mRNA expression levels of *DLST* in different clinical subgroups, including T stage, N stage, M stage, pathologic stage, and histologic grade.

**Table 1:**
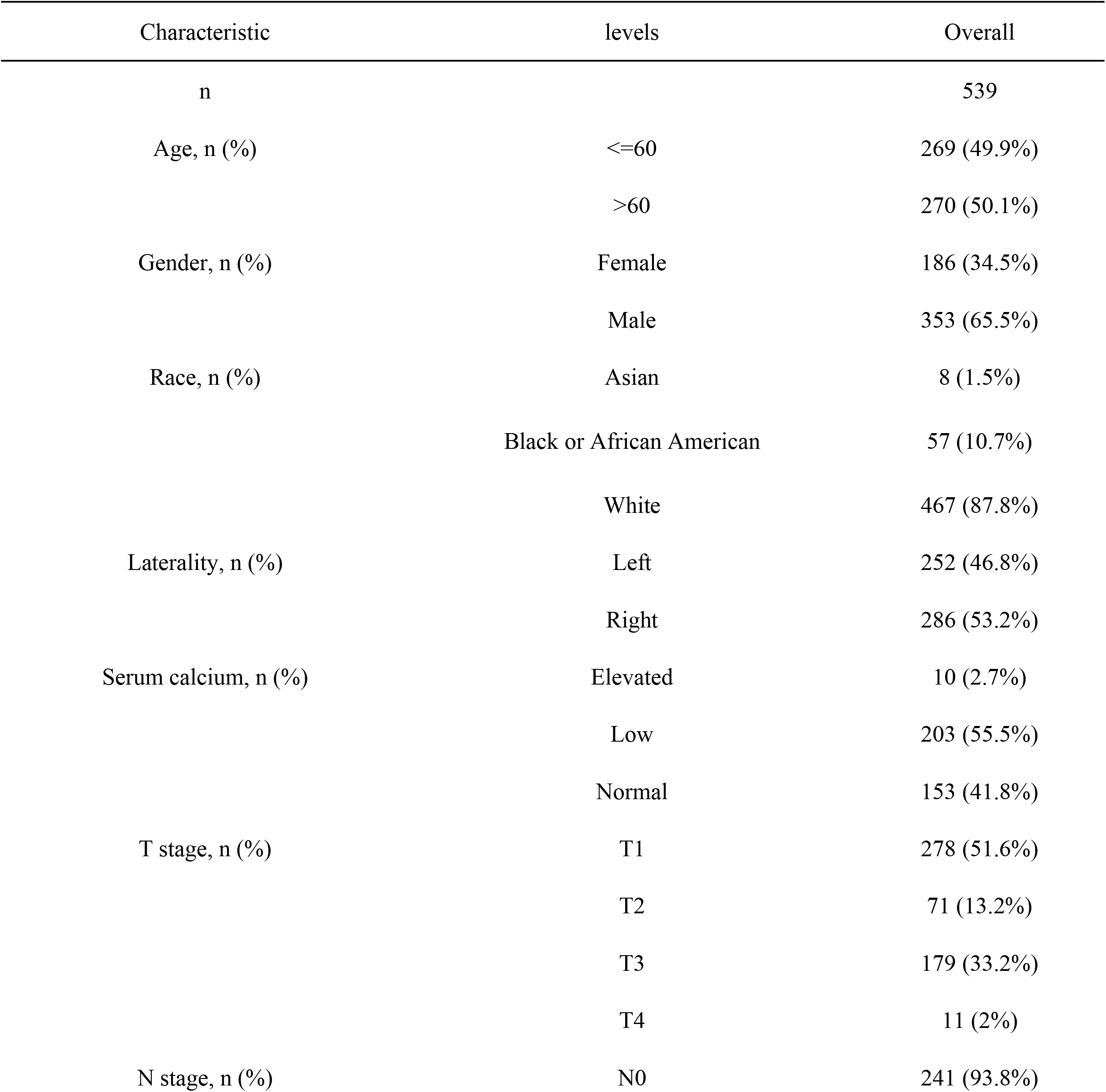
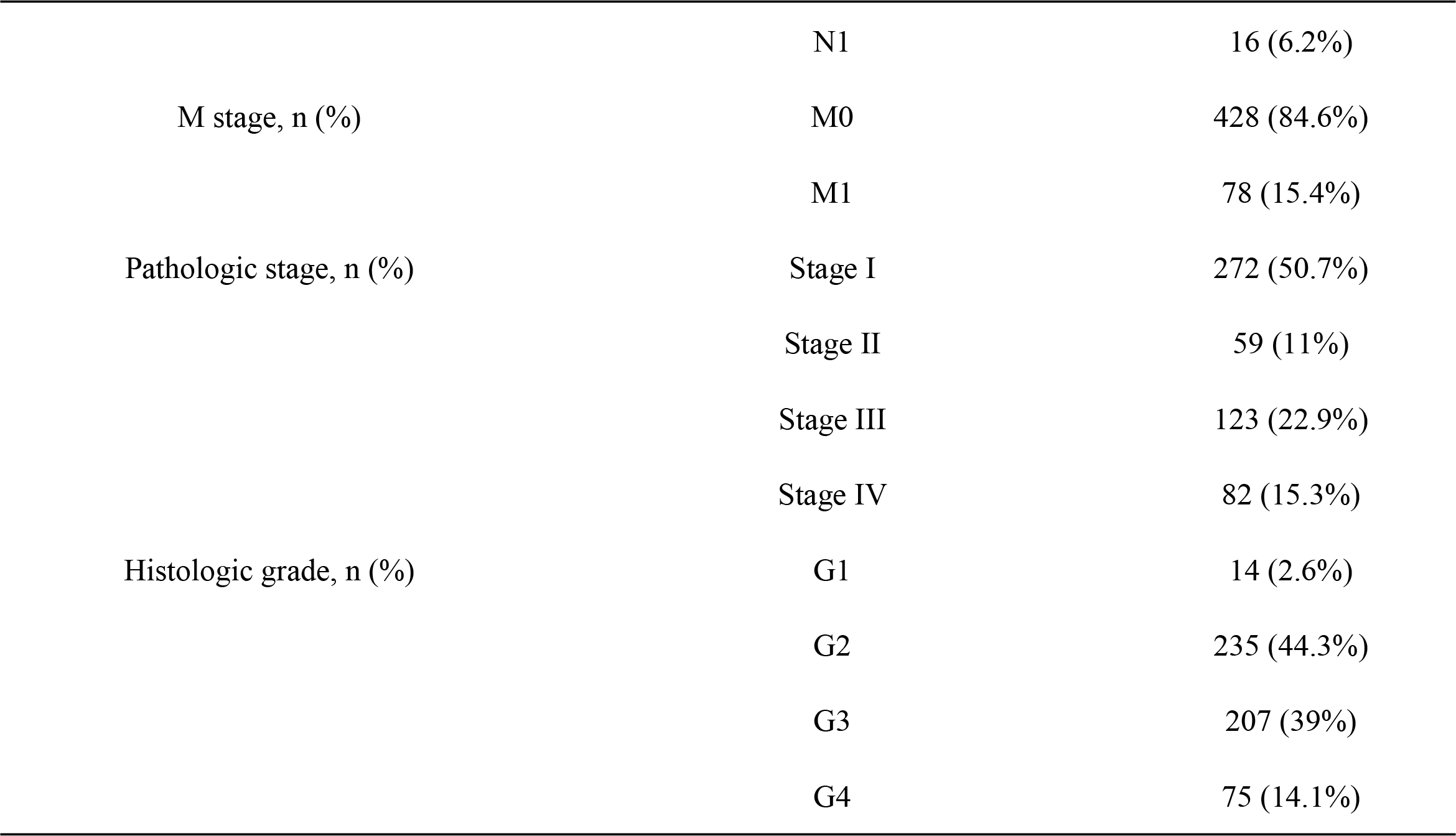
Demographic data and clinical information for all samples.

### 2.5 Functional enrichment and gene set enrichment analysis (GSEA) of differentially expressed genes (DEGs) between samples with high and low *DLST* expression levels

The median *DLST* expression was regarded as the cut-off value to identify DEGs between the two groups (low and high expression) of *DLST* in TCGA data. The “DESeq2” R package was used for analysis, and the “ggplot2” R package was used to construct the volcano plot. The threshold of DEGs for functional enrichment analysis was defined as |logFC| > 2 and an adjusted p-value < 0.05. The GSEA method identifies gene or protein classes that are overrepresented in a large set of genes or proteins. The predefined gene set was from the MSigDB database (http://www.gsea-msigdb.org/gsea/msigdb/index.jsp). GO and Kyoto Encyclopedia of Genes and Genomes (KEGG) analyses were performed using the “ClusteProfiler” R package to explore the functional and pathway differences between the two groups with differential *DLST* expression. For each analysis, the permutation number was set to 1000. Enrichment results that met the conditions of adjusted p-value < 0.05 and false discovery rate-adjusted q-value < 0.25 were defined as statistically significant.

### 2.6 Statistical analysis

Based on the median expression of *DLST*, the high and low expression levels were determined. We analyzed the association between *DLST* expression and clinicopathological parameters using the Pearson method. Comparative analyses of clinical and pathological characteristics between the high- and low-expression groups were performed using the chi-square test. Univariate Cox proportional hazards regression was used to estimate the individual HRs for OS. Cox proportional hazard regression was used to identify independent prognostic factors in ccRCC patients. A nomogram and a prediction model were constructed in R (version 3.6.3). Statistical analyses were conducted using SPSS software (version 22, IBM, SPSS, Chicago, IL, USA). Significant results are indicated by p < 0.05, and highly significant results are indicated by p < 0.01.

## 3 Results

### 3.1 Differential expression of CRGs in ccRCC samples

We compared the expression of eight CRGs in normal tissue and tumor samples, using TCGA and GTEx data. The expression of *DLST, FDX1, PDHX, COX7B*, and *NDUFB1* was significantly downregulated in ccRCC samples, whereas the expression of *SLC6A3, CCS*, and *MID1* was significantly upregulated, compared to the expression in the control samples (Figure 1A). The genes were ranked according to the intensity and significance of their differential expression (Figure 1B). The expression of *FDX1, DLST, COX7B, NDUFB1*, and *PDHX* was higher than the expression of *CCS, MID1*, and *SLC6A3*, according to the log2 FC. The expression of the eight genes was externally validated using the GEO database (GSE78179 and GSE53757), and five genes (*DLST, NDUFB1, PDHX, COX7B*, and *CCS*) were also found to be expressed in the samples in the GEO datasets, which was in agreement with the results from TCGA (Figure 1C).

**Figure 1:**
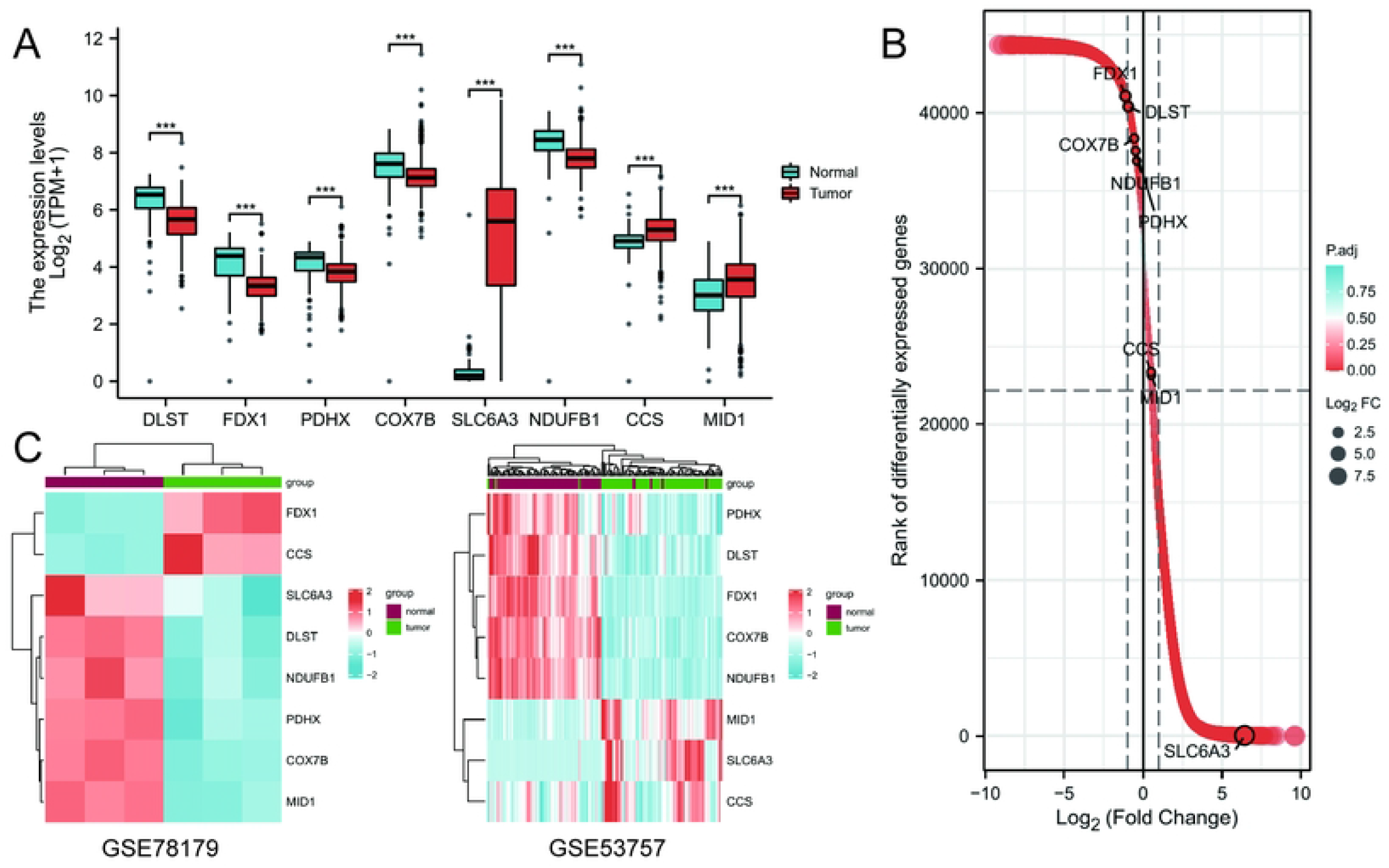
The expression of eight CRGs between normal tissues and ccRCC in TCGA and GEO datasets. ^***^p<0.001. (A) Differential expression between normal tissues and ccRCC samples in TCGA; (B) The location of the eight CRGs according to the rank of differential expression genes in ccRCC; (C) Verification of differential expression in GEO datasets

### 3.2 Gene–gene and protein–protein interactions among the CRGs

Based on TCGA data, we further investigated the expression of the CRGs, and the heat map in Figure 2A shows that the eight CRGs were highly correlated with each other. The expression of *DLST* was highly positively correlated with the expression of *FDX1, PDHX, SLC6A3* and *NDUFB1*. Next, we analyzed the functional interaction network at the gene and protein levels, using the GeneMANIA network (Figure 2B). The results revealed that gene–gene interactions accounted for 54.45% of all functions, and the predicted value was 19.81%. The protein–protein interaction network revealed that *DLST* and *NDUFB1*, which exhibited extensive connections to other genes, were the hub genes in the network (Figure 2C).

**Figure 2:**
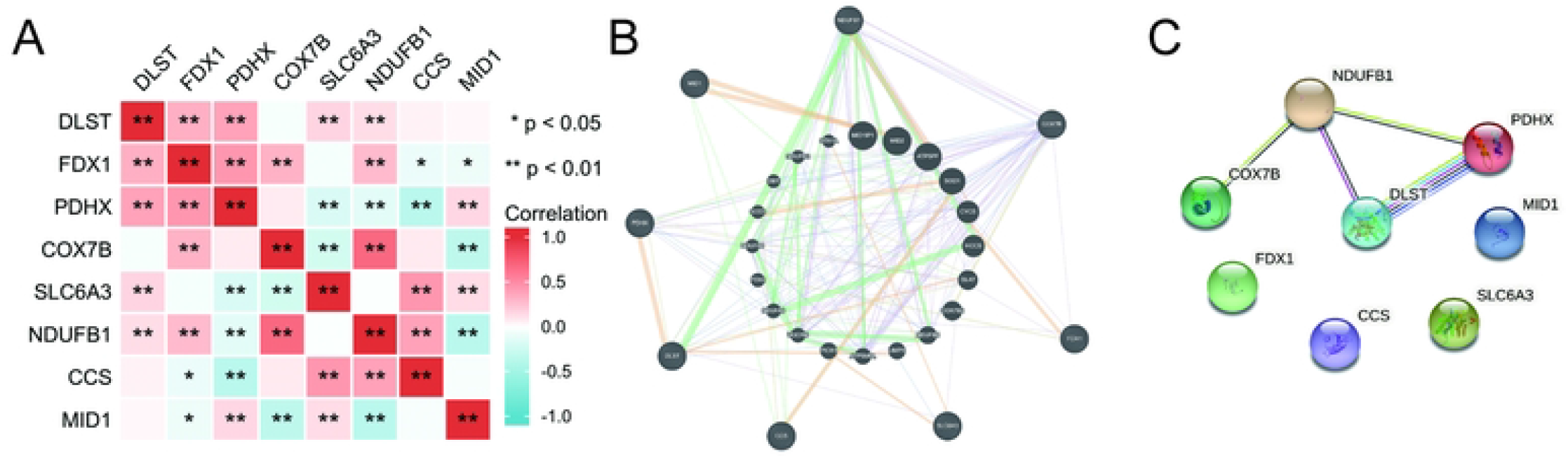
Correlation analysis of the eight CRGs. ^*^p<0.05; ^**^p<0.01. (A) Correlation heatmap of the eight CRGs; (B) The network of eight CRGs interacting with target genes; (C) Protein–protein interaction network of eight CRGs

### 3.3 Diagnostic and prognostic values of eight CRGs

The expression of the eight CRGs was significantly different in the 72 ccRCC samples, compared to that in the matched adjacent samples (Figure 3A). Thus, we evaluated the diagnostic and prognostic roles of these CRGs in ccRCC patients. We used ROC curves to test whether these genes could be used for diagnosis; genes with area under the ROC curve (AUC) above 0.85 were considered more accurate for diagnosing diseases. Five genes (*DLST, FDX1, PDHX, COX7B*, and *SLC6A3*) exhibited a relatively high diagnostic value for ccRCC (Figure 3B). Using Kaplan–Meier survival curves, we revealed that the low expression of *DLST* had a significant prognostic value (p < 0.01) (Figure 3C, D). Further experiments were conducted on *DLST* because of its highly significant prognostic value.

**Figure 3:**
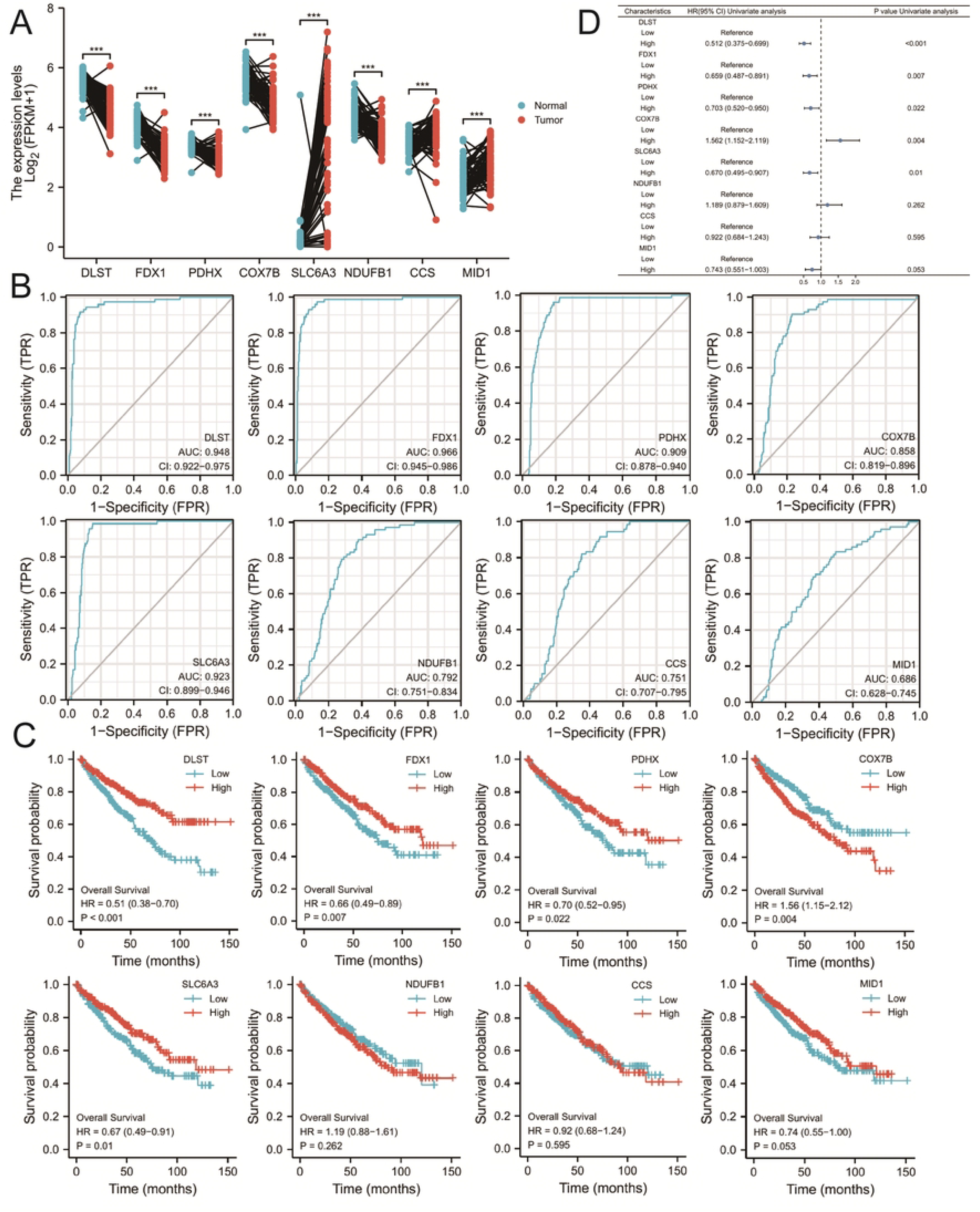
Prognostic and diagnostic value analysis. ^***^p<0.001. (A) The differential expression of eight CRGs in paired samples; (B) ROC curve for eight genes in adjacent TCGA-KIRC tissues and TCGA-KIRC samples; (C) Kaplan–Meier overall survival analyses of eight CRGs in ccRCC; (D) Forest plot with univariate Cox regression analysis of the expression level of eight genes

### 3.4 Low expression of *DLST* is correlated with clinicopathological features of ccRCC patients

Compared to that in the normal group, the expression of *DLST* in the ccRCC group was significantly decreased (p < 0.01). *DLST* mRNA expression differed significantly according to TNM stage, pathologic stage, and histological grade. The analysis revealed that *DLST* expression was significantly downregulated in T3 and T4 stage, N1 stage, M1 stage, pathologic stage III and IV, and histological grade 3 and 4 ccRCC patients (p < 0.01) (Figure 4A). Immunohistochemical staining analysis revealed that *DLST* expression was lower in the ccRCC group than in the normal tissue group (Figure 4B). Low expression of *DLST* may promote tumor invasion, metastasis, and malignancy. Thus, the patients were divided into high- and low-*DLST* expression groups, based on the mean *DLST* expression level (Table 2). In the low-expression group, the ratios of the clinical features associated with high degree of malignancy were significantly higher than those in the high-expression group.

**Table 2:** The relationship between *DLST* expression and clinical characteristics in ccRCC.

| Characteristic | levels | Low expression of *DLST* | High expression of *DLST* | p |
| --- | --- | --- | --- | --- |
| n |  | 269 | 270 |  |
| Age, n (%) | <=60 | 132 (24.5%) | 137 (25.4%) | 0.763 |
|  | >60 | 137 (25.4%) | 133 (24.7%) |  |
| Gender, n (%) | Female | 77 (14.3%) | 109 (20.2%) | 0.005 |
|  | Male | 192 (35.6%) | 161 (29.9%) |  |
| Race, n (%) | Asian | 3 (0.6%) | 5 (0.9%) | 0.329 |
|  | Black or African American | 24 (4.5%) | 33 (6.2%) |  |
|  | White | 239 (44.9%) | 228 (42.9%) |  |
| Laterality, n (%) | Left | 122 (22.7%) | 130 (24.2%) | 0.600 |
|  | Right | 146 (27.1%) | 140 (26%) |  |
| Serum calcium, n (%) | Elevated | 7 (1.9%) | 3 (0.8%) | 0.209 |
|  | Low | 94 (25.7%) | 109 (29.8%) |  |
|  | Normal | 81 (22.1%) | 72 (19.7%) |  |
| T stage, n (%) | T1 | 113 (21%) | 165 (30.6%) | < 0.001 |
|  | T2 | 40 (7.4%) | 31 (5.8%) |  |
|  | T3 | 111 (20.6%) | 68 (12.6%) |  |
|  | T4 | 5 (0.9%) | 6 (1.1%) |  |
| N stage, n (%) | N0 | 120 (46.7%) | 121 (47.1%) | 0.008 |
|  | N1 | 14 (5.4%) | 2 (0.8%) |  |
| M stage, n (%) | M0 | 201 (39.7%) | 227 (44.9%) | 0.001 |
|  | M1 | 53 (10.5%) | 25 (4.9%) |  |
| Pathologic stage, n (%) | Stage I | 108 (20.1%) | 164 (30.6%) | < 0.001 |
|  | Stage II | 30 (5.6%) | 29 (5.4%) |  |
|  | Stage III | 75 (14%) | 48 (9%) |  |
|  | Stage IV | 54 (10.1%) | 28 (5.2%) |  |
| Histologic grade, n (%) | G1 | 1 (0.2%) | 13 (2.4%) | < 0.001 |
|  | G2 | 94 (17.7%) | 141 (26.6%) |  |
|  | G3 | 117 (22%) | 90 (16.9%) |  |
|  | G4 | 56 (10.5%) | 19 (3.6%) |  |
| OS event, n (%) | Alive | 158 (29.3%) | 208 (38.6%) | < 0.001 |
|  | Dead | 111 (20.6%) | 62 (11.5%) |  |
| DSS event, n (%) | Alive | 188 (35.6%) | 232 (43.9%) | < 0.001 |
|  | Dead | 76 (14.4%) | 32 (6.1%) |  |
| PFI event, n (%) | Alive | 162 (30.1%) | 216 (40.1%) | < 0.001 |
|  | Dead | 107 (19.9%) | 54 (10%) |  |

**Figure 4:**
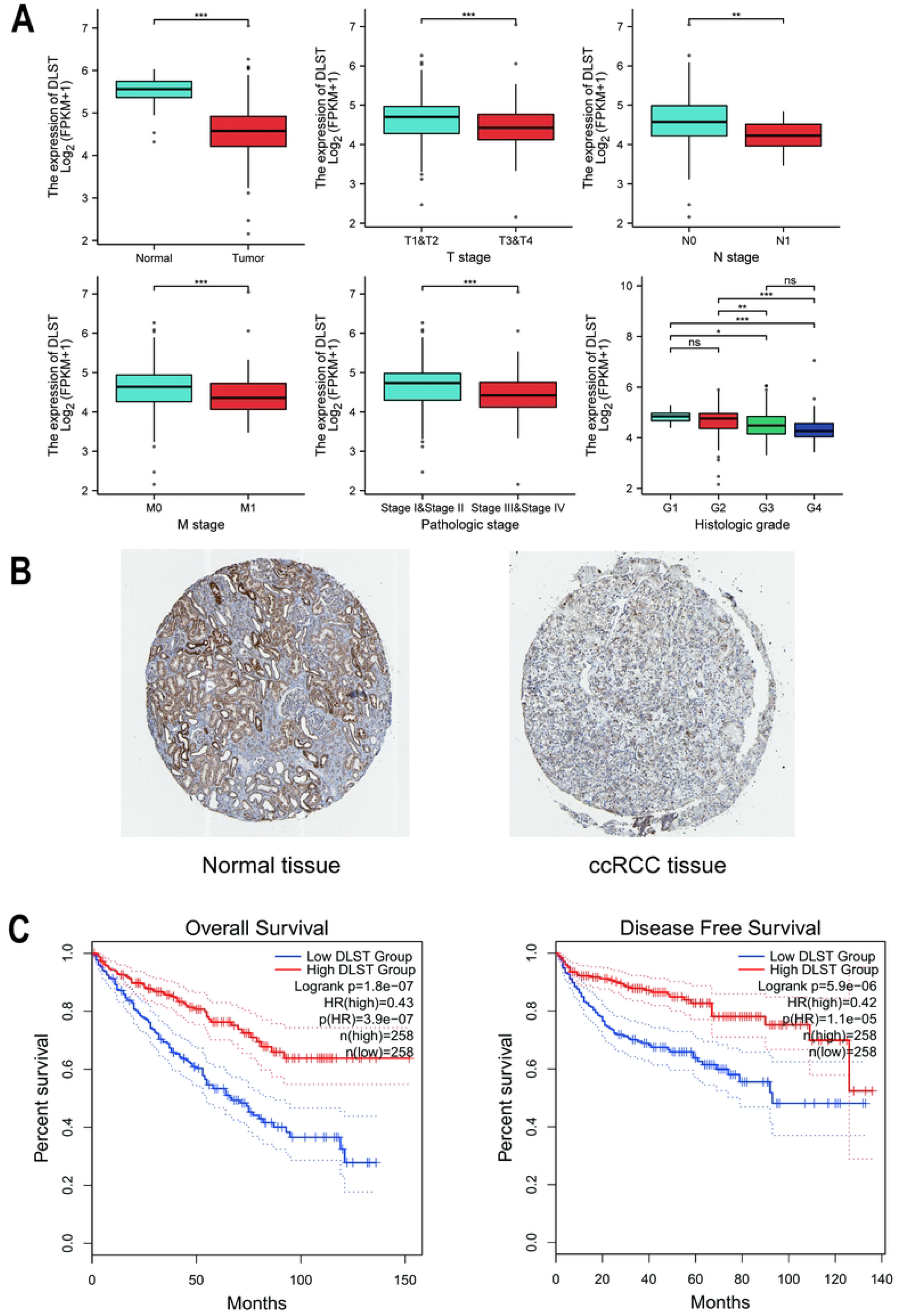
Expression of DLST and its clinical value. (A) Association between *DLST* expression and clinical features. ns, p>0.05; ^*^p<0.05; ^**^p<0.01; ^***^p<0.001. (B) Immunohistochemical staining of DLST in normal tissues and ccRCC; (C) Kaplan– Meier overall survival analyses and disease-free survival analyses of *DLST*

### 3.5 Relationship between *DLST* expression and ccRCC prognosis

To elucidate the relationship between low *DLST* expression levels and patient survival, survival analyses were performed using GEPIA2. Low *DLST* expression was found to correlate with poor OS (p = 3.9e-07) and DFS (p = 1.1e-05) (Figure 4C).

Potential predictors, including age, sex, T stage, M stage, pathologic stage, and *DLST* expression level, were examined using Cox regression analyses. Univariate analysis revealed significant associations between OS and age, T stage, M stage, pathologic stage, and *DLST* expression level (p < 0.001 for all; Table 3). These risk factors were further studied using a multivariate Cox regression analysis, which revealed that *DLST* could be a potential independent prognostic factor (HR = 0.665, 95% CI = 0.470–0.941, p = 0.021). The nomograms were modeled based on the clinical features and the expression level of *DLST* in ccRCC patients. The results showed that the higher the score, the worse the prognosis. The predicted probabilities of the calibration plots were consistent with the observed results (Figure 5).

**Table 3:**
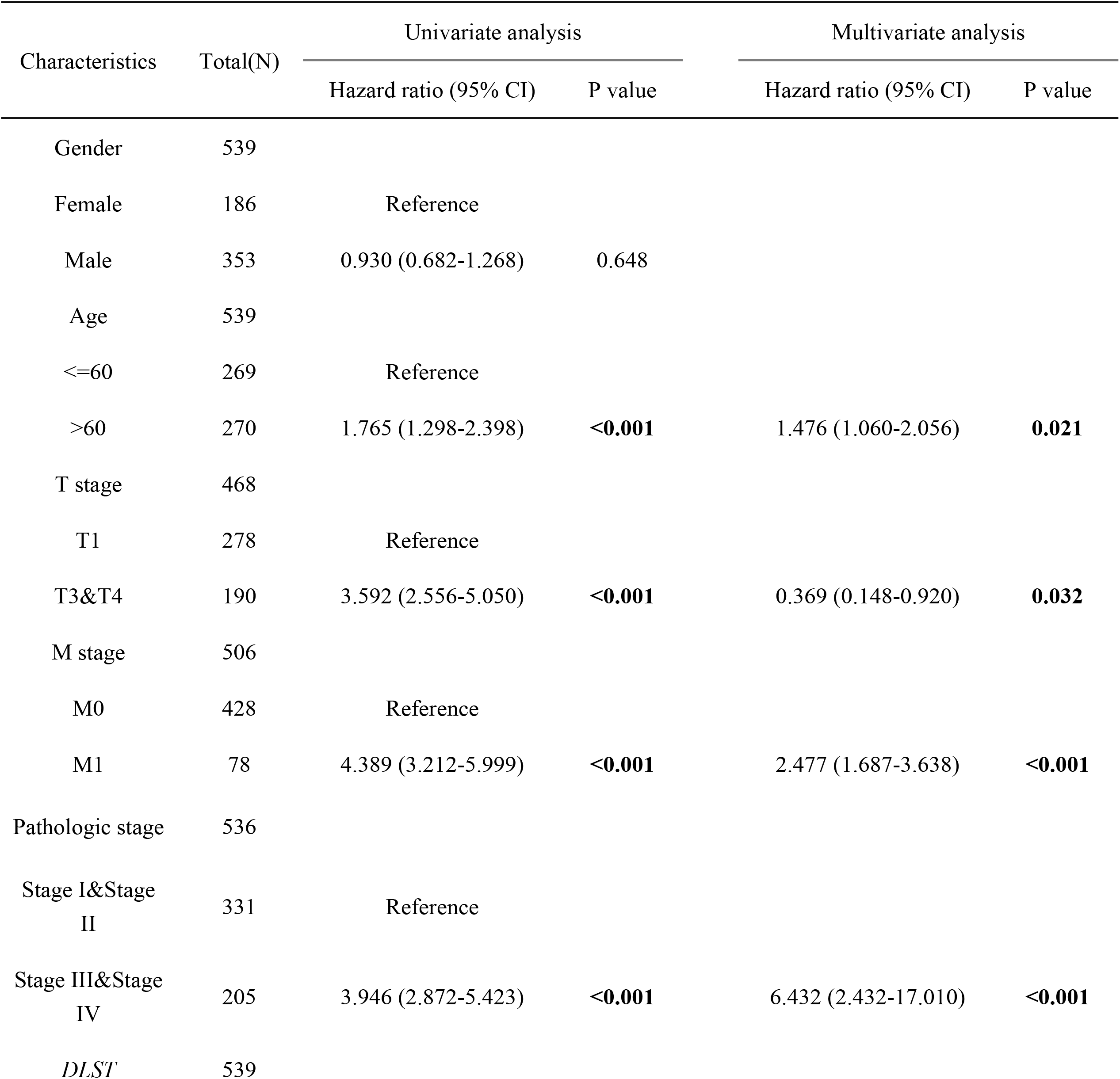
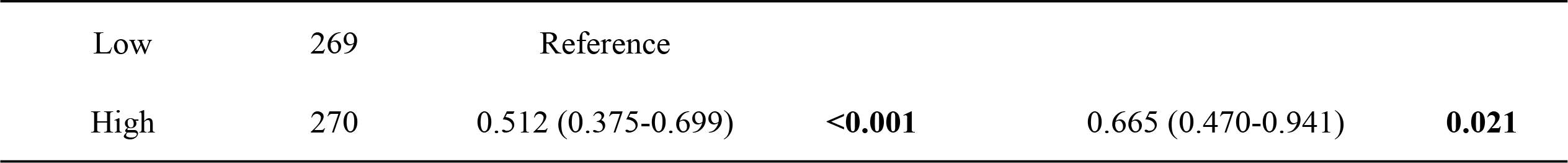
Univariate Cox regression and multivariate Cox regression analysis of OS in ccRCC.

| Characteristics | Total(N) | Univariate analysis |  | Multivariate analysis |  |
| --- | --- | --- | --- | --- | --- |
|  |  | Hazard ratio (95% CI) | P value | Hazard ratio (95% CI) | P value |
| Gender | 539 |  |  |  |  |
| Female | 186 | Reference |  |  |  |
| Male | 353 | 0.930 (0.682-1.268) | 0.648 |  |  |
| Age | 539 |  |  |  |  |
| <=60 | 269 | Reference |  |  |  |
| >60 | 270 | 1.765 (1.298-2.398) | <0.001 | 1.476 (1.060-2.056) | 0.021 |
| T stage | 468 |  |  |  |  |
| T1 | 278 | Reference |  |  |  |
| T3&T4 | 190 | 3.592 (2.556-5.050) | <0.001 | 0.369 (0.148-0.920) | 0.032 |
| M stage | 506 |  |  |  |  |
| M0 | 428 | Reference |  |  |  |
| M1 | 78 | 4.389 (3.212-5.999) | <0.001 | 2.477 (1.687-3.638) | <0.001 |
| Pathologic stage | 536 |  |  |  |  |
| Stage I&Stage II | 331 | Reference |  |  |  |
| Stage III&Stage IV | 205 | 3.946 (2.872-5.423) | <0.001 | 6.432 (2.432-17.010) | <0.001 |
| DLST | 539 |  |  |  |  |
|  |  | Hazard ratio (95% CI) | P value | Hazard ratio (95% CI) | P value |
| Low | 269 | Reference |  |  |  |
| High | 270 | 0.512 (0.375-0.699) | **<0.001** | 0.665 (0.470-0.941) | **0.021** |

**Figure 5:**
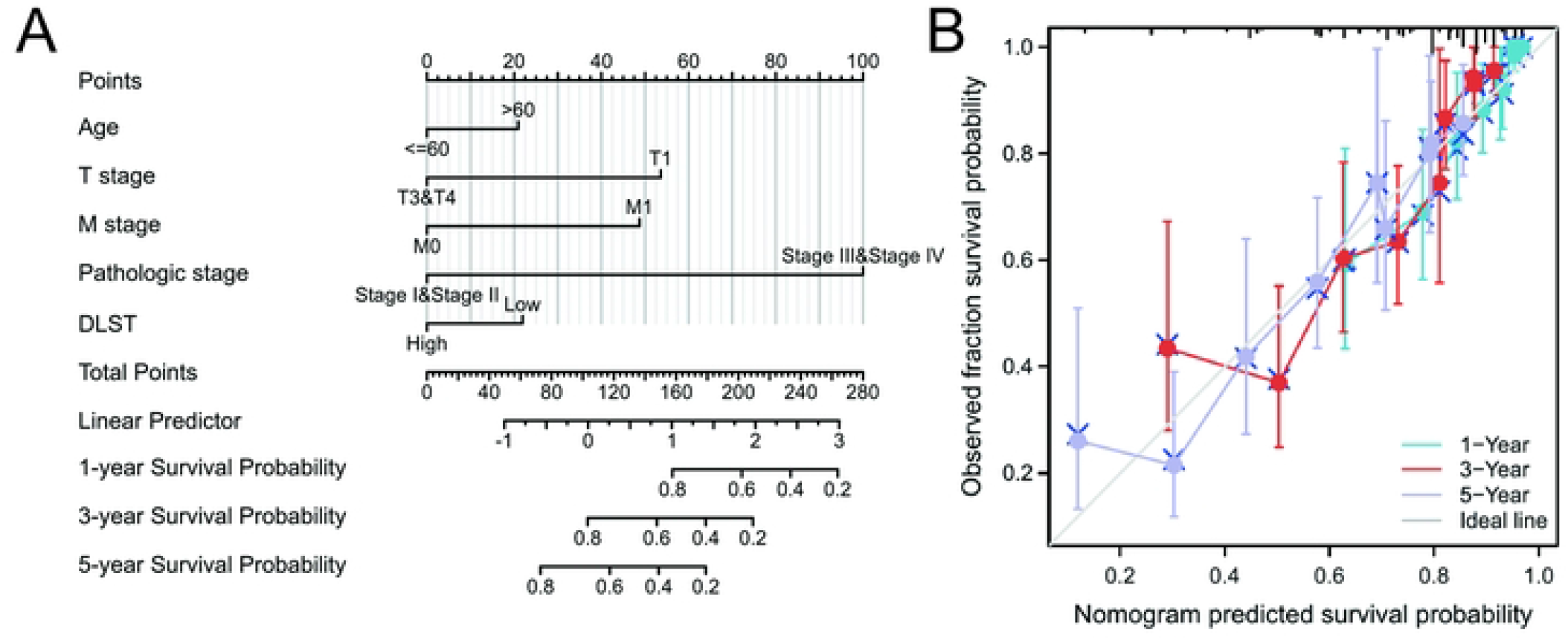
Prognostic prediction model of *DLST* in ccRCC. (A) Nomogram for 1-year, 3-year, and 5-year OS of ccRCC patients; (B) Calibration plots for 1-year, 3-year, and 5-year OS prediction

### 3.6 Functional enrichment of DEGs in differential *DLST* expression level groups

DEGs were identified between the two *DLST* expression groups (low and high expression) based on |logFC > 1| and adjusted p-value = 0.05 (Figure 6A). GO analyses (Figure 6) revealed that DEGs were enriched in the following biological process (BP) terms: complement activation, classical pathway, humoral immune response mediated by circulation immunoglobulin, and protein activation cascade; in the following cellular component (CC) terms: immunoglobulin complex, immunoglobulin complex, circulating microparticles, and blood microparticles; and in the following molecular function (MF) terms: antigen binding and receptor ligand activities. KEGG analysis revealed that the DEG was enriched in the following pathways: complement and coagulation cascades, systemic lupus erythematosus, and neuroactive ligand–receptor interactions. Figures 6D and 6E show that GO:00019814, GO:0006985, GO:0002455, and GO:0072376 had higher Z-scores, indicating that the downregulation of *DLST* may be associated with the inhibition of immune-related functions and with the development of ccRCC.

**Figure 6:**
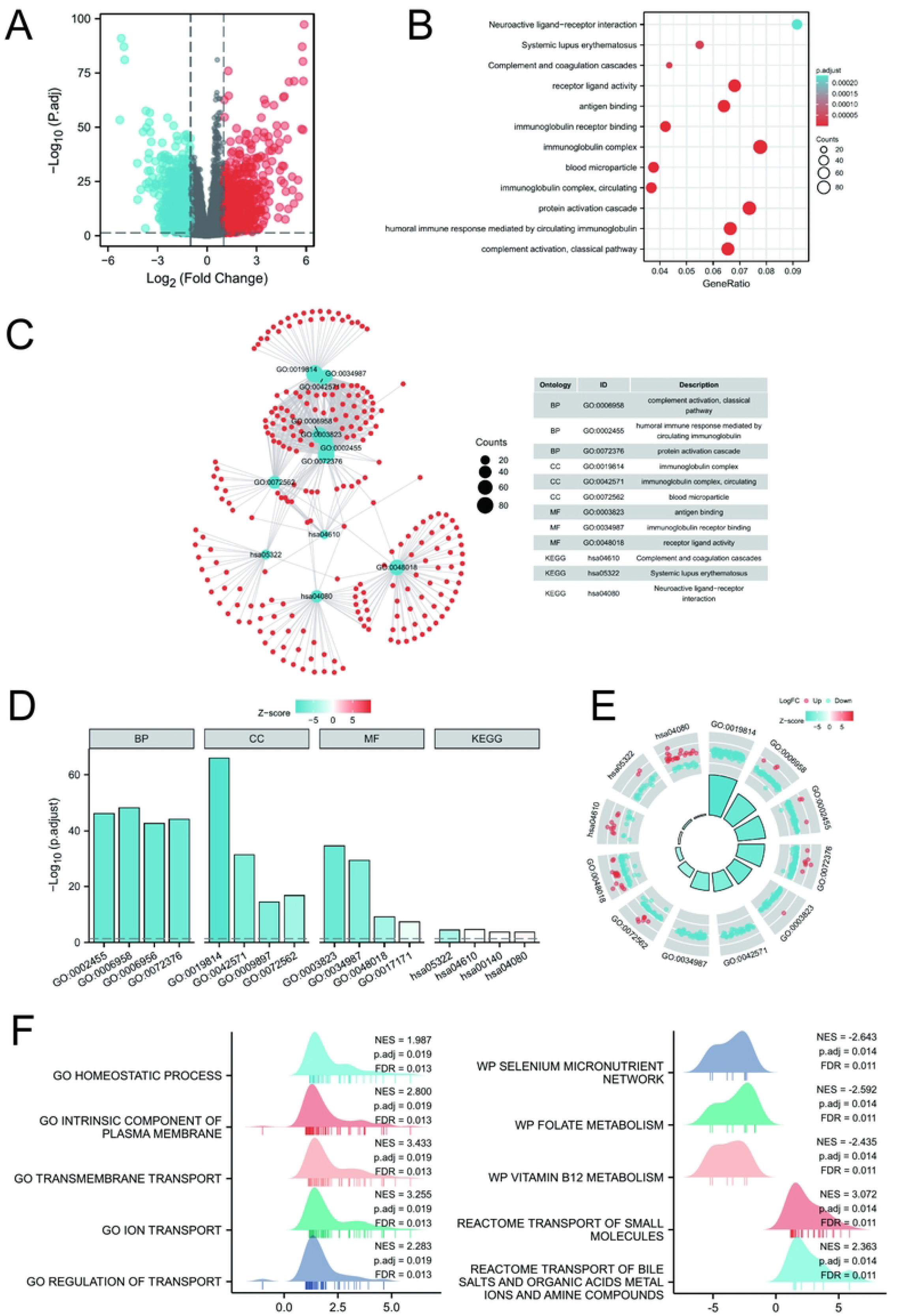
Functional enrichment analysis based on differential genes between low- and high-*DLST* expression groups. (A) Volcanogram of differential genes between low- and high-*DLST* expression groups; (B) Bubble diagram of GO and KEGG enrichment; (C) Clustered molecular networks of GO enrichment analysis (BP: biological process, CC: cellular component, MF: molecular function) and KEGG pathway annotation; (D) Bar graph after combining logFC and GO/KEGG functional clustering; (E) Loop graph after combining logFC and GO/KEGG functional clustering; (F) Enrichment analyses from GSEA

We performed GSEA to further identify the biological functions involved in ccRCC samples with different *DLST* expression levels (Figure 6F). The results showed that the DEGs were enriched in GO homeostatic process, GO intrinsic component of plasma membrane, GO transmembrane transport, GO ion transport, GO regulation of transport, WikiPathways (WP) selenium micronutrient network, WP folate metabolism, WP vitamin B12 metabolism, reactome transport of small molecules, and reactome transport of bile salts and organic acids, metal ions, and amine compounds.

## 4 Discussion

In this study, we examined the expression of eight CRGs for their potential diagnostic and prognostic utility in ccRCC. Four genes (*DLST, FDX1, PDHX*, and *SLC6A3*) had good diagnostic efficacy (AUC > 0.9), and five (*DLST, FDX1, PDHX, COX7B*, and *SLC6A3*) were associated with prognosis. Gene expression levels were analyzed using TCGA data, and the expression was verified using two datasets downloaded from the GEO database. The expression of *DLST* was significantly downregulated in all three datasets. *DLTA* has been identified as a gene that confers resistance against cuproptosis. *DLST* is a paralog of *DLTA*; thus, *DLST* could be a vital gene related to cuproptosis and ccRCC. However, the upstream and downstream molecular mechanisms of *DLST* in cuproptosis require further investigation.

*DLST* is a protein-coding gene (Wang et al. 2017), and Remacha et al. (2019) found that it is associated with paragangliomas and hereditary paraganglioma-pheochromocytoma syndromes. The pathways associated with *DLST* include glucose/energy metabolism and TCA cycle III. Shen et al. (2021) found that the triple-negative breast cancer cell lines (Hs578T and SUM158PT) had relatively lower DLST expression at both the transcript and protein levels. *DLST* inactivation significantly impacted multiple metabolic pathways, such as aminoacyl-tRNA biosynthesis, pyrimidine metabolism, amino-acid metabolism pathways, and the TCA cycle, in DLST-dependent cell lines. Buffet et al. (2020) suggested that *DLST* is one of the susceptibility genes of paragangliomas and pheochromocytomas, which encodes one of the three components (the E2 component) of the 2-oxoglutarate dehydrogenase complex that catalyzes the overall conversion of 2-oxoglutarate to succinyl-CoA and CO(2). Moreover, Anderson et al. (2021) revealed that *DLST* loss significantly suppressed NADH production and impaired oxidative phosphorylation, and that the high expression of *DLST* predicts poor treatment outcome and aggressive disease in neuroblastoma patients. These previous studies suggest that *DLST* plays an important role in a variety of tumors. Some studies suggest that high expression of DLST indicates poor prognosis, but this study showed that low expression of DLST led to poor prognosis in ccRCC. The reason for this discrepancy may lie in the different types of energy metabolism in different primary tumor sites. In the process of ccRCC metabolism, oxidative phosphorylation level decreases, fatty acid synthesis accumulates, and aerobic glycolysis and lipid metabolism occupy the main metabolic position, which may induce *DLST* to play an important role in ccRCC. This phenomenon may be due to the relatively more important role of lipid metabolism in ccRCC.

Analysis of *DLST* expression in different clinical subgroups, such as T3 and T4 stage, N1 stage, M1 stage, pathologic stage III and IV, and histological grade 3 and 4 patients, showed that its expression was downregulated in patients with poor prognosis. Our data indicate that *DLST* might play a critical role in ccRCC initiation and progression through immunological-related mechanisms and cuproptosis. To improve the accuracy of identifying high-risk patients, we constructed a prognostic nomogram that involved age, T stage, M stage, pathologic stage, and *DLST* expression levels as indicators. In addition, the Kaplan–Meier survival curves showed that patients with low *DLST* expression had poor OS and DFS.

Gene function enrichment revealed that the DEGs in the low- and high-expression *DLST* groups were related to systemic immune function. In the low-expression group, systemic immune functions were inhibited. The result showed that downregulation of *DLST* resulted in the downregulation of immune-related genes and changes in immune regulatory functions. For example, when the expression of *DLST* was low, the expression of immunoglobulin complexes was downregulated, which may affect the occurrence and development of tumors. However, studies focused on investigating the relationship between *DLST* and tumor immune function are currently very limited. *DLST* may be involved in the synthesis and regulation of related immune proteins in ccRCC, but this still requires further mechanistic studies for confirmation.

This study has some limitations. Further experimental evidence is needed to demonstrate the mechanism of *DLST* in ccRCC and the changes in cuproptosis function in the groups with low *DLST* expression.

The eight CRGs (*DLST, FDX1, PDHX, COX7B, SLC6A3, NDUFB1, CCS*, and *MID1*) were differentially expressed in normal and ccRCC patients, and only *DLST* was found to be a potential diagnostic and prognostic factor in patients with ccRCC. *DLST* might be a potential predictor to guide clinical practice.

## Data Availability

All RNA sequence profiling files are available from the TCGA database

https://portal.gdc.cancer.gov/

## 5 Conflict of Interest

The authors declare that the research was conducted in the absence of any commercial or financial relationships that could be construed as a potential conflict of interest.

## 6 Author Contributions

HW and XN designed the study. XM and HW performed the statistical analysis. SL and XM drafted the manuscript. All authors read and approved the final manuscript.

## 7 Funding

The study was founded by Department of Finance of Hebei China [grant no. jcyf (2020)397].

## 8 List of abbreviations

AUC: Areas under the ROC curve
BP: Biological process
CC: Cellular component
ccRCC: Clear cell renal cell carcinoma
CCS: Copper chaperone for superoxide
CRGs: Cuproptosis-related genes
DEGs: Differentially expressed genes
DFS: Disease-free survival
DLST: Dihydrolipoamide S-succinyltransferase
FC: Fold change
GEO: Gene Expression Omnibus
GO: Gene Ontology
GSEA: Gene set enrichment analysis
HR: Hazard ratio
KEGG: Kyoto Encyclopedia of Genes and Genomes
MF: Molecular function
OS: Overall survival
RCC: Renal cell carcinoma
ROC: Receiver operating characteristic
TCA: Tricarboxylic acid
TCGA: The Cancer Genome Atlas

## 9 Acknowledgments

Not available.

## 10 Third party support

None third party support was received for this study.

## 12 Supplementary Material

Not available.

## 13 Data Availability Statement

The datasets analyzed for this study can be found in the TCGA (https://portal.gdc.cancer.gov/), GEO (https://www.ncbi.nlm.nih.gov/geo/query/acc.cgi?acc=GSE78179 and https://www.ncbi.nlm.nih.gov/geo/query/acc.cgi?acc=GSE53757), UCSC Xena (https://xenabrowser.net/hub/), and the Human Protein Atlas (www.proteinatlas.org).

## Notes

### Competing Interest Statement

The authors have declared no competing interest.

### Funding Statement

The author(s) received no specific funding for this work.

